# Experience in the validation of a rapid test for NS1 and IgM for early diagnosis during a dengue epidemic in Colombia

**DOI:** 10.1101/2023.11.08.23298285

**Authors:** Rosa-Margarita Gélvez Ramírez, Mónika Patricia Consuegra, María Isabel Estupiñan, Adriana Torres Rangel, Víctor Mauricio Herrera, Xavier de Lamballerie, Luis Ángel Villar Centeno

## Abstract

Dengue has a broad spectrum of syndromic presentations, making clinical diagnosis difficult in regions where acute febrile syndrome has multiple etiologies. Rapid tests for detecting NS1 and IgM are broadly proposed for the early diagnosis of dengue; however, their implementation in primary care settings is inconsistent, and the relevance of IgM detection in this context has not been firmly established. Our study aimed to describe the experience of validating an NS1-IgM rapid test in primary care settings in Bucaramanga, Colombia, during a dengue epidemic (2018 to 2020). We tested blood samples from 568 patients with a clinical diagnosis of dengue using the Bioline Dengue Duo rapid test and RT-PCR as a reference to estimate sensitivity, specificity, and positive and negative predictive values (SE, SP, PPV, and NPV, respectively). The prevalence of infection was 32.9% (95%CI: 29.1, 36.8), whereas SE and SP were 83.4% and 72.7% for NS1 without statistical heterogeneity across disease duration. NS1’s PPV and NPV were 60.0% and 89.9% at the observed prevalence. Our results show that NS1 and IgM rapid tests (POCT) are feasible in the primary care setting. The contribution of the NS1 test was indisputable, with high-performance levels far superior to those of the IgM test. The NS1+IgM combination did not offer a significant advantage over NS1 alone.

## Introduction

Dengue, a vector-borne arboviral infection, is currently one of the diseases with the greatest health burden among populations with active transmission, showing a steadily increasing trend in the number of cases during the last decade and transitioning from endemicity to hyperendemicity probably due to a lack of effective surveillance, vector control, and diagnostic strategies (1). In 2019, the number of infected individuals, hospitalizations, and fatal cases increased ten-fold compared to the previous year (2). In the Americas, 2,809,818 cases were reported in 2022, with a cumulative incidence of 283 per 100,000 inhabitants, and up to the 10^th^ epidemiological week of 2023, dengue accounted for about 3 out of 4 cases of arboviral infections (3).

In Colombia, the epidemiological behaviour of dengue is complex due in part to the presence of competent vectors such as *Aedes aegypti* and *Aedes albopictus*, the circulation of all four serotypes of the dengue virus (DENV), the immunological susceptibility of the population, a long history of internal migration and more recently, the phenomena of massive migration across borders (4). During the last two decades the country has witnessed outbreaks in 1998, 2002, 2010, 2013, and 2019, with case-fatality rates above 2% (4). Throughout 2023 the Colombian Ministry of Health has reported 56,585 cases of dengue, including 821 severe cases, which represents an approximately nine-fold increase compared to the same period in 2022 (5). The east-central region of the country has been historically one of the most affected areas by dengue, recording 24,997, 49,059, 45,726, and 46,598 cases in the epidemics of 2002, 2010, 2013, and 2019, respectively (6).

In dengue endemic areas of Latin-American the differential diagnosis for acute fever illness comprises a broad spectrum of infections including another arbovirus. A major challenge in clinical diagnosis is to differentiate dengue from other infectious diseases such as chikungunya, Zika, leptospirosis, influenza, and even HIV (7–10). For instance, dengue and chikungunya have overlapping symptoms such as fever, headache, rash, and arthralgia during the first days of disease, which potentially leads to misdiagnosis (11). Notably, a proportion of dengue cases will progress to a life-threatening severe disease; however, prognostic factors, collectively known as warning dengue signs (WDS), are nonspecific and often develop late in the course of illness (12). In this context, the access to an early and accurate diagnosis of dengue infection, particularly in the primary healthcare setting, would improve the opportunity to stratify risk and closely monitor patients with high likelihood of progression to complications and death.

According to the Centers for Disease Control and Prevention (CDC), a positive nucleic acid amplification test (NAAT) or a positive antigenic test (nonstructural protein 1; NS1) performed in a single sample obtained during the acute phase of disease (≤7 days after fever onset) is considered a confirmatory diagnosis for dengue (13). Testing for IgM antibodies is recommended as a complementary approach during the acute phase; however, it is most useful when performed in convalescent samples (>7 days post symptom onset) although a positive result in a single specimen is only considered presumptive of dengue virus infection. Unfortunately, due to the shortage in infrastructure, supplies, and trained staff, it is not possible to perform NAATs in most of the Colombian health centers, and even IgM antibody detection tests usually require referral of samples to mass-processing reference laboratories. These limitations preclude any attempt to make early etiological diagnosis of dengue, scenario in which medical management of patients almost exclusively relies on clinical diagnosis.

In this context, the development and implementation of point-of-care testing (POCT) strategies, whose results are obtained in real-time, that is, in parallel to the clinical evaluation of patients, constitute a promising alternative to overcome the barriers of etiological diagnosis in resource-limited environments. These strategies should comply with the WHO’s requirements according to the ASSURED criteria: Affordable, Sensitive, Specific, User-friendly, Rapid and Robust, Equipment-free, and Deliverable to end-users (14–17). In the case of dengue, POCT technologies, specifically rapid diagnostic tests (RDT), have shown to be highly reliable, although heterogeneous in terms of their diagnostic accuracy compared to NAATs (18–20). The aim of this report is to describe the experience around the validation of a RDT to detect NS1 antigens and IgM antibodies as a POCT in a primary healthcare facility for the early diagnosis of dengue during an epidemic in Colombia.

## Methods

In 2018, the World Health Organization (WHO) and the Pan American Health Organization (PAHO) issued an epidemiological alert considering “the onset of the season of increased dengue transmission in the Southern Hemisphere” and recommended that “Member States implement preparedness and response actions to prevent dengue transmission and avoid deaths from this disease” (21). In response to this alert, a complementary surveillance system was established in the context of the public health network of Bucaramanga, Colombia, between 2018 and 2020, with the purpose of improving early detection of dengue cases during the acute phase of the disease. As part of this initiative, an immunochromatography test (rapid test-POCT) for the detection of DENV non-structural protein 1 (NS1) and IgM antibodies against DENV was implemented to assist the early diagnosis of febrile patients.

### Study area

This study was conducted in Bucaramanga, a midsize city located in the northeast of Colombia. This city has a public health network named “Instituto de Salud de Bucaramanga (ISABU)” composed by 22 primary health-care facilities and 2 teaching hospitals: the “Unidad Materno Infantil Santa Teresita (UIMIST) and the “Hospital Local del Norte (HLN)” that provide first level of complexity services.

### Eligibility and definition of clinically suspected dengue cases

Patients who consecutively sought medical care at the HLN between December 2018 and July 2019 were eligible for the study. In addition, patients had to be classified as clinically suspected cases of dengue, defined as a history of febrile illness of 1 to 5 days’ duration and at least one of the following symptoms or signs: headache, arthralgia, myalgia, retro orbital pain, positive tourniquet test, petechiae, or hemorrhagic manifestations. A representative sample of cases presenting a diagnostic dilemma underwent clinical examination by a trained physician and provided a blood sample for a complete blood count and diagnostic tests.

### Diagnostic testing

First, the remaining blood samples after the complete blood count were analyzed in the laboratory of the HLN by a trained microbiologist, blinded to clinical information, using the Bioline Dengue Duo (NS1 Ag + IgM/IgG; Abbott Ref 11FK46) according to the manufacturer’s recommendations. Bioline Dengue Duo is an immunochromatographic qualitative test (rapid test with negative/positive results) that allows the simultaneous detection of NS1 antigen and antibodies class IgM and IgG against DENV.

As temporary storage after the rapid tests, the remaining samples were centrifuged for 10 min at 3.500 rpm, aliquoted and stored at -80°C in the biorepository. All sample aliquots, regardless of the results of the rapid tests and blind of it, were sent to the reference laboratory Unité des Virus Émergents (UVE) at Aix-Marseille University in France. This was justified because in the pre-pandemic era of COVID-19 we did not have the infrastructure, reagents and equipment necessary to perform these tests. Once at UVE, the samples were thawed and trained personnel performed the tests using the molecular biology real-time PCR (RT-PCR) test for the genomic detection of DENV, CHIKV and ZIKV, considered the reference test. In case of positive results, serotyping tests for DENV-1, DENV-2, DENV-3, and DENV-4 were performed.

RNA extraction was performed using the commercial kit QIAamp Viral RNA Mini QIAcube Kit (QIAGEN 52926) by automated method. Once the RNA was purified, retrotranscription and amplification were performed in a single step using the commercial enzyme SuperScript™ III Platinum™ One-Step qRT-PCR (11732088) and the primer and probe amplification systems, provided by the EVAg (European Virus Archive-GLOBAL), following a previously published amplification protocol (22,23); a positive RT-PCR result was considered with a CT (Cycle Threshold) of ≤37. Those who interpreted the rapid test and the RT-PCR reference standard were blinded to the other result.

An aliquot of these samples were processed locally for viral isolation. C6/36 HT cells were inoculated with samples diluted 1:25 in 1 mL of L-15 medium supplemented with 2% fetal bovine serum, penicillin-streptomycin and L-glutamine. Cells were incubated at +34°C and observed daily for the presence of cytopathic effect (CPE). On day 7, or earlier if CPE was detected, the supernatant was collected and cells were fixed for processing by indirect immunofluorescence (IFI).

### Ethical Considerations

This study was approved by the Scientific Technical Committee of the HLN-ISABU, the Institutional Review Board and Ethical Committee of the “Centro de Atención y Diagnóstico de Enfermedades Infecciosas (CDI)” in Bucaramanga and the Ministry of Health and Social Protection of Colombia.

### Statistical analysis

We estimated the sensitivity and specificity of the NS1 rapid test contrasted against RT-PCR along with their corresponding exact binomial 95% confidence intervals (95%CI) for the whole sample and stratifying by disease duration. We contrasted accuracy indexes by estimating relative probabilities (RP; i.e., the ratio of sensitivities [RPse] or specificities [RPsp]) using generalized linear models (logarithmic link function) separately, according to the reference result (24): RPs for sensitivities and specificities were estimated in patients with positive and negative RT-PCR results, respectively. We estimated the positive (PPV) and negative (NPV) predictive values for a predefined set of prevalences ranging from 10-50%, using Bayes’ theorem (25): First, we calculated post-test odds of disease as the product of pre-test odds (from the assumed prevalences) and the diagnostic likelihood ratios (DRLs) of positive and negative test results, respectively. Then, we transformed the positive and negative post-test odds into their respective predictive values. This procedure was repeated for each assumed prevalence by disease duration while updating the DRLs accordingly. We derived 95%CI for the predictive values by estimating first the lower and upper 95% confidence limits of the DLRs (26) and then replacing those values into the calculation described above, following the Bayesian approach.

## Results

Our cohort was conducted in the context of the complementary surveillance system as we describe above. We included 568 patients who sought medical care at the HLN where the cases were evaluated sequentially between December 2018 and July 2019. The median age of participants was 13 years (interquartile range: 6 – 25 years; 62.7% were children), and 269 (47.4%) were male. Table 1 shows the distribution of positive results from NS1 and IgM rapid tests, independently and combined, and RT-PCR by disease duration. Overall, the prevalence of infection as defined by a positive RT-PCR was 32.9% (IC95%: 29.1, 36.8), similar to that based on a positive IgM rapid test but significantly lower than that based on a positive NS1 rapid test: 33.1% and 45.8%, respectively. On the other hand, 54.0% of samples were defined as positive when considering the combination of NS1+IgM results.

**Table 1.** RT-PCR and rapid tests positivity (%) by disease duration.

| Duration | n | Test |  |  |  |
| --- | --- | --- | --- | --- | --- |
|  |  | RT-PCR | NS1 | IgM | NS1+IgM* |
| 0 | 15 | 60.0 (32.3, 83.7) | 73.3 (44.9, 92.2) | 40.0 (16.3, 67.7) | 73.3 (44.9, 92.2) |
| 1 | 38 | 26.3 (13.4, 43.1) | 39.5 (24.0, 56.6) | 18.4 (7.7, 34.3) | 42.1 (26.3, 59.2) |
| 2 | 34 | 38.2 (22.2, 56.4) | 35.3 (19.7, 53.5) | 14.7 (5.0, 31.1) | 41.2 (24.6, 59.3) |
| 3 | 95 | 41.1 (31.1, 51.6) | 45.3 (35.0, 55.8) | 24.2 (16.0, 34.1) | 49.5 (39.1, 59.9) |
| 4 | 126 | 53.2 (44.1, 62.1) | 55.6 (46.4, 64.4) | 27.8 (20.2, 36.5) | 63.5 (54.4, 71.9) |
| 5 | 121 | 21.5 (14.5, 29.9) | 48.8 (39.6, 58.0) | 47.1 (38.0, 56.4) | 61.2 (51.9, 71.9) |
| 6 | 53 | 18.9 (9.4, 32.0) | 41.5 (28.1, 55.9) | 43.4 (29.8, 57.7) | 50.9 (36.8, 64.9) |
| 7 | 37 | 16.2 (6.2, 32.0) | 35.1 (20.2, 52.5) | 40.5 (24.8, 57.9) | 45.9 (29.5, 63.1) |
| 8 | 23 | 21.7 (7.5, 43.7) | 43.5 (23.2, 65.5) | 39.1 (19.7, 61.5) | 52.2 (30.6, 73.2) |
| 9 | 10 | 20.0 (2.5, 55.6) | 50.0 (18.7, 81.3) | 50.0 (18.7, 81.3) | 60.0 (26.2, 87.8) |
| 10 | 6 | 0.0 (0.0, 45.9) | 0.0 (0.0, 45.9) | 33.3 (4.3, 77.7) | 33.3 (4.3, 77.7) |
| ≥11 | 10 | 0.0 (0.0, 30.9) | 0.0 (0.0, 30.9) | 9.5 (0.2, 44.5) | 10.0 (0.3, 44.5) |
| **All** | **568** | **32.9 (29.1, 37.0)** | **45.8 (41.6, 50.0)** | **33.1 (29.2, 37.1)** | **54.0 (49.9, 58.2)** |

\* Positive/negative combined result: NS1[+] or IgM[+] / NS1[-] and IgM[-].

Regarding disease duration, 429 (75.5%) and 519 (89.9%) patients provided samples within the first 5- and 7-days post-onset of symptoms: positivity to RT-PCR and NS1 rapid test was higher in samples of patients with ≤5 versus >5 days of disease duration (38.2% versus 16.6%, and 48.9% versus 35.9%, respectively), as well as in samples of patients with ≤7 versus >7 days of disease duration (34.7% versus 14.3%, and 47.2% versus 30.6%, respectively). The opposite occurred in the case of IgM, particularly comparing samples collected from patients with ≤5 versus >5 days of disease duration: 31.0% versus 39.6%, respectively (Table 1, Figure 1).

**Figure 1.**
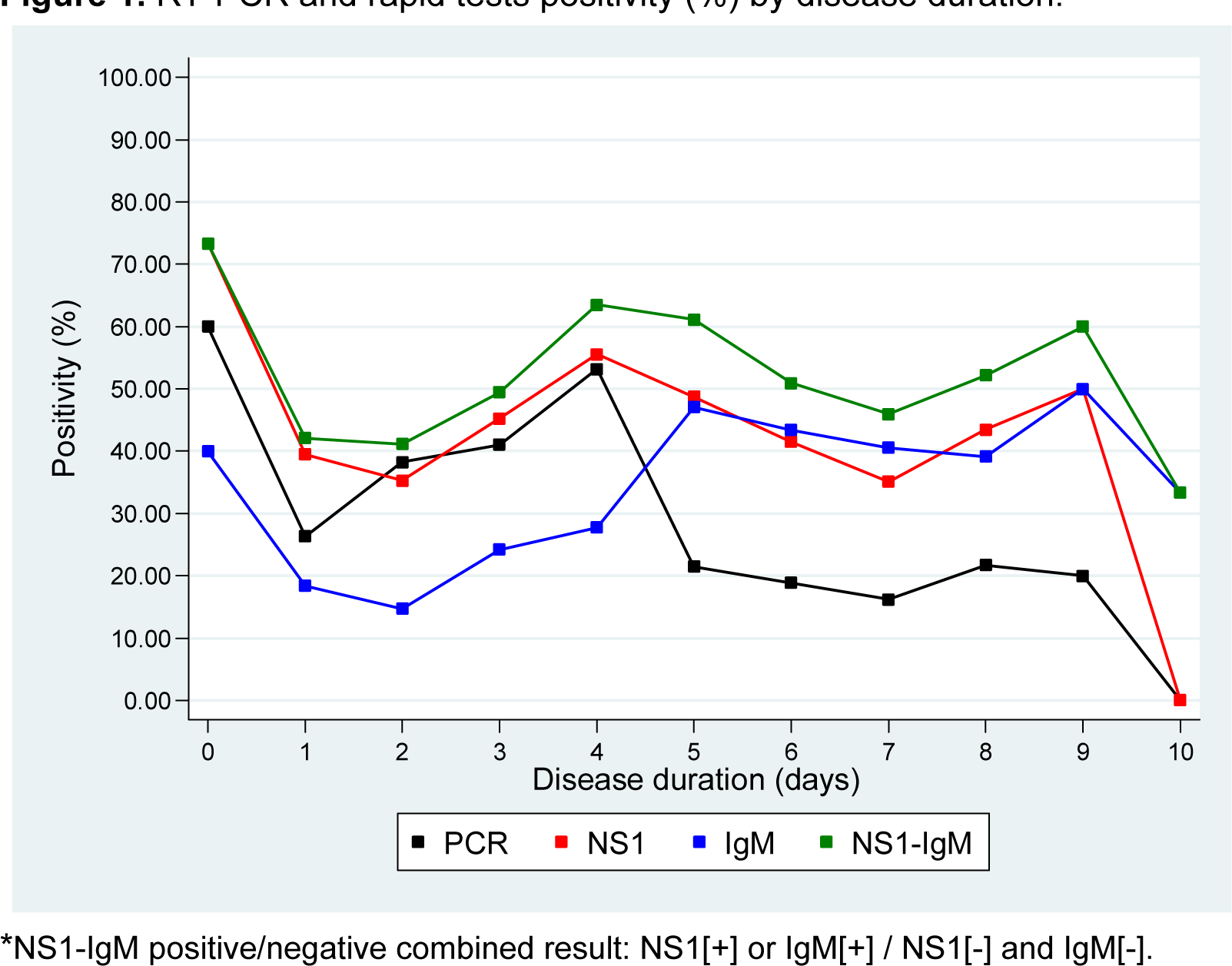
RT-PCR and rapid tests positivity (%) by disease duration.

The overall sensitivity and specificity of NS1 were 83.4% (95%CI: 77.3, 88.5) and 72.7% (95%CI: 67.9, 77.1), respectively; however, when evaluated up to 5- and 7-days post-onset of symptoms the corresponding estimates were 84.2% (95%CI: 77.6, 89.4) and 72.8% (95%CI: 67.0, 78.1), and 83.9% (95%CI: 77.7, 88.9) and 72.3% (95%CI: 67.2, 77.0), respectively. Neither sensitivity nor specificity differed statistically across disease duration (Table 2, Figure 2). Evaluated at the observed prevalence of infection (RT-PCR positivity of 32.9%), the PPV and NPV of the NS1 rapid test were 60.0% (95%CI: 53.8, 66.0) and 89.9% (95%CI: 86.0, 93.1), however as expected, values fluctuate according to the day of illness and the prevalence of the disease. The highest value corresponds to day 2 being for PPV 100% for all the prevalence and for NPV 99% to a prevalence of 10% (95%CI: 94.6, 98.8) (Figure 3), when sensitivity and specificity had their highest values.

**Table 2.** Sensitivity, specificity, and relative probabilities (RP) of NS1 by disease duration up to 7 days from disease’s onset.

| Duration | n | Sensitivity | RP* | Specificity | RP* |
| --- | --- | --- | --- | --- | --- |
| 0 | 15 | 88.9 (51.8, 99.7) | 1.00 | 50.0 (11.8, 88.2) | 1.00 |
| 1 | 38 | 90.0 (55.5, 99.7) | 1.01 (0.74, 1.38) | 78.6 (59.0, 91.7) | 1.57 (0.69, 3.58) |
| 2 | 34 | 92.3 (64.0, 99.8) | 1.04 (0.79, 1.37) | 100.0 (83.9, 100.0) | 2.09 (0.94, 4.66) |
| 3 | 95 | 82.1 (66.5, 92.5) | 0.92 (0.70, 1.21) | 80.4 (67.6, 89.8) | 1.61 (0.71, 3.61) |
| 4 | 126 | 82.1 (70.8, 90.4) | 0.92 (0.71, 1.19) | 74.6 (61.6, 85.0) | 1.49 (0.66, 3.37) |
| 5 | 121 | 84.6 (65.1, 95.6) | 0.95 (0.72, 1.26) | 61.1 (50.5, 70.9) | 1.22 (0.54, 2.76) |
| 6 | 53 | 70.0 (34.8, 93.3) | 0.78 (0.49, 1.26) | 65.1 (49.1, 79.0) | 1.30 (0.57, 2.99) |
| 7 | 37 | 100.0 (54.1, 100.0) | 1.18 (0.94, 1.48) | 77.4 (58.9, 90.4) | 1.55 (0.68, 3.52) |

\* RP: Relative probability (references: 0 days for duration).

**Figure 2.**
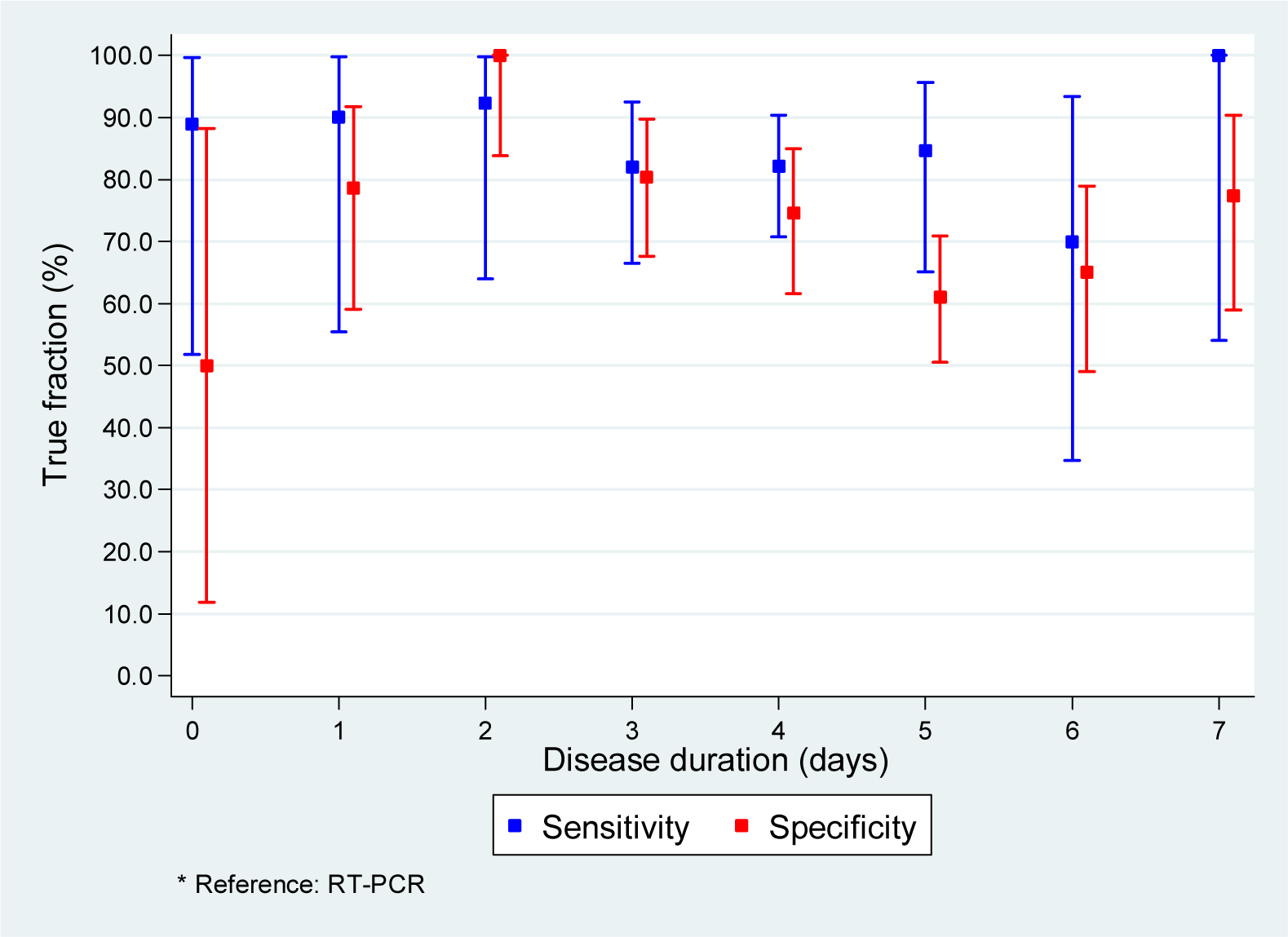
Sensitivity and specificity of NS1 rapid test, compared against RT-PCR*, by disease duration up to 7 days from disease’s onset.

**Figure 3.**
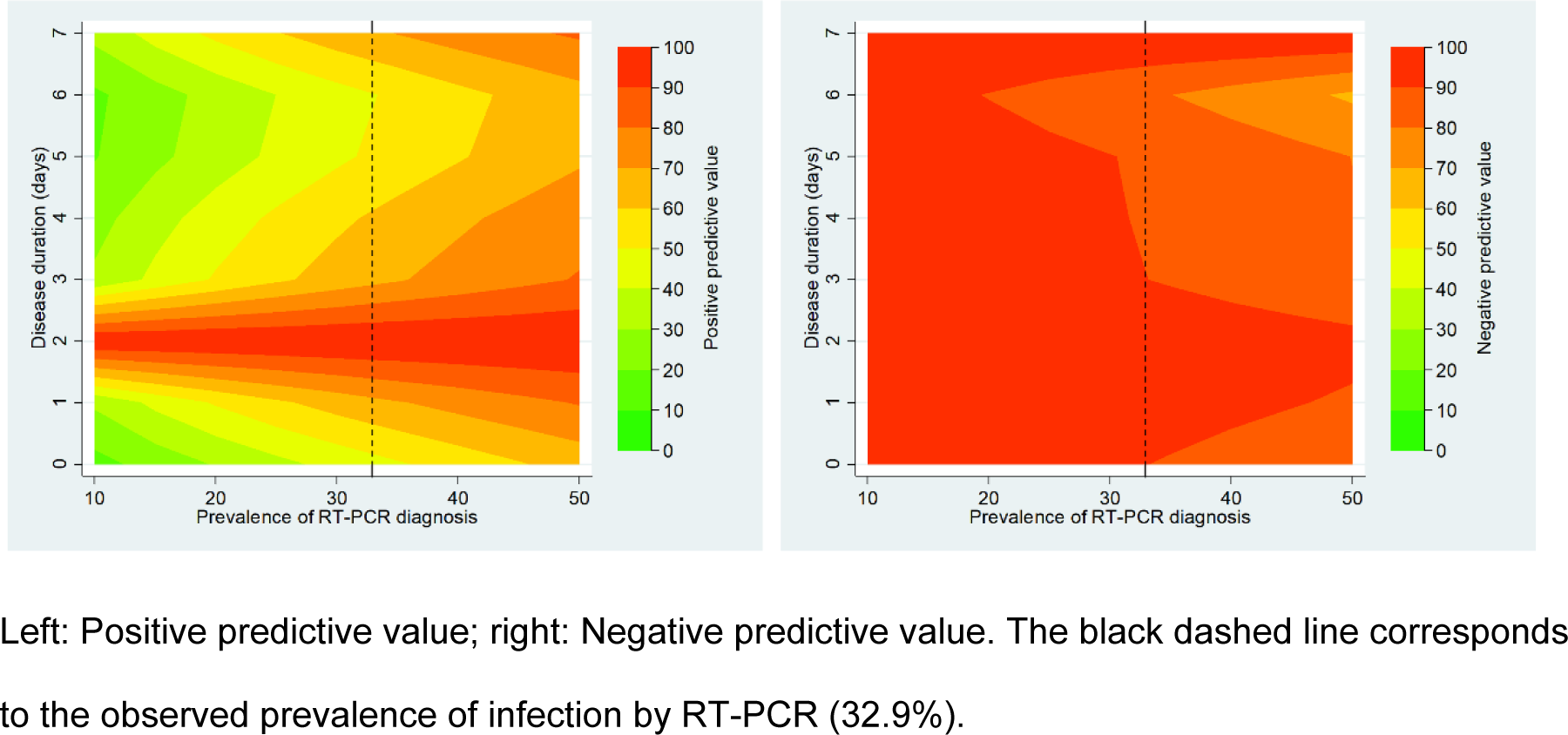
Predictive values of NS1 rapid test by disease duration and a set of assumed prevalences.

All samples were processed by a molecular biology test for genomic detection of DENV, CHIKV and ZIKV (Trioplex) in the same reaction. In these samples, only the DENV genome was detected and for CHIKV and ZIKV the results were negative. From all samples with a positive NS1 result on the rapid test were obtained virus isolation. Of the positive cases for DENV, the infecting serotype was determined, with 96% corresponding to DENV-1 and 4% to DENV-2. Virus isolation was achieved for each of these serotypes, yielding information amenable to sequencing tests for future phylogeny and phylogeography studies

## Discussion

After the introduction of CHIKV and ZIKV, during 2014 and 2015 respectively, a new DENV epidemic occurred during the years 2018 to 2020 in Colombia (27). During this period, this study was carried out with the purpose of implementing in a primary care center in the city of Bucaramanga, the use of a rapid immunochromatographic test to diagnose dengue disease in an area where the nonspecific febrile illness is multi-etiological. This study showed that the NS1+IgM rapid test is not only feasible in the context of primary care at the same point where patients are seen, but also proved to be an accurate method for the diagnosis of dengue in endemic areas. This suggesting that it is possible to make an early and adequate diagnosis at the patient’s first visit (28–33).

During the study period, in our country dengue rapid tests were not authorized for laboratory-confirmed diagnosis of dengue disease, so virological or serological reference tests should always be applied (34). This implied transferring the samples to other reference laboratories with the specific capacity (RT-PCR test) or collecting a second sample in the early convalescent phase of disease in order to be able to perform serological tests to detect IgM seroconversion. This circuit meant that the opportunity for results in less than 24 hours was extended to 96 hours or even weeks. This was a late diagnosis and, in situations of outbreaks and epidemics, the physician would initiate clinical management according to the WHO guidelines (12) without the availability of confirmatory laboratory tests. Follow-up to obtain a second sample was difficult since patients did not return to the health center when they felt better (35). This situation created a culture for many years in which patients only attended their health center when they already felt very ill, even with warning dengue signs-WDS. Thus, when they arrived at the primary care health center, some cases required a fast referral to more complex health-care services. The purpose of the introduction of this rapid test was offered to physicians a tool for the reduction of their clinical uncertain during the first seventh days of onset of dengue illness and for patients to understand the importance of this 89.9% (n=519) of the patients attended and provided specimens during this acute phase of the disease. The prevalence of dengue in the population study was 32.9%. According to reports from the Instituto Nacional de Salud-INS and the Ministerio de Salud y Protección Social, the dengue in Colombia in ER 01 to 07 was placed in a situation of alert. Since ER 08 presented a behavior above the expected number of cases at the national level, compared to its historical behavior (2011–2018), which placed the country in an epidemic situation for week 52 of 2019 (36). For this same period in the Americas region PAHO/WHO reported that the epidemic, which had spread in the region by 2019, was contributing a significant percentage of the population under 15 years of age (37). In our findings, the median age of the patients corresponded to 13 years (interquartile range: 6 - 25 years) where 62.7% were children. These results are therefore in accordance with the epidemiology by age group affected during this epidemic in the Americas region. Studies have shown that the greatest susceptibility to DENV infections corresponds to young patients where primary infections prevail in this age group and the estimated risk of dengue increases with age (38).

Regarding the percentage of positivity, this corresponded to 45.8% for NS1 and 33.1% for IgM. If we assume that 89.9% of the patients were in the acute phase of the disease, it is reasonable to think that the greater proportion of positives corresponds to the detection of NS1 antigen and not IgM antibodies against DENV. However, when we combine the percentage of positivity for NS1 and IgM this value increases to 54% positivity. According to the Centers for Disease Control and Prevention (CDC) recommendations for the diagnosis of dengue, a negative RT-PCR or NS1 result cannot rule out the diagnosis. On the contrary, it is recommended that during the first 7 days of illness, molecular biology tests such as RT-PCR, NS1 antigen detection and IgM antibody detection be combined to increase case detection in a single sample during the acute phase of the disease (13). The results of this study are in agreement with this recommendation, since it is possible to observe an increase of almost 10% in the detection of dengue cases when combining NS1 and IgM tests in the same sample during the first 7 days of illness compared to only detecting NS1 or IgM separately. Having a device that can detect both NS1 and IgM at the same time could allow the detection window to be extended during a longer phase of the disease and that the performance advantages of both NS1 and IgM complement each other in the diagnosis of DENV.

The results discriminated by day of illness are also consistent; in the acute phase of the disease versus the convalescent phase (≤7 versus >7 days) the highest positivity is given by RT-PCR and NS1 with 34.7% versus 14.3%, and 47.2% versus 30.6%, respectively. The opposite occurs with IgM where its positivity increases during the convalescent phase of the disease. This reiterates the concept that DENV diagnostic tests are adjusted to the natural history of the disease, where direct detection of the virus (RNA or antigens) is done in the early phases of the disease where viral replication is active and indirect detection tests (IgM antibodies against DENV) occur in the convalescent phases where the immune response begins to develop (39,40). This antibody response and its detection will also depend on the type of infection the patient is suffering from; for primary infections, IgM is detected from the fifth day of illness and IgG from the tenth day of illness. On the contrary, in secondary infections, the IgG present from the previous infection increases rapidly and IgM could be detected at the same time as IgM (40,41). The type of infection was not evaluated in order to see the effect on sensitivity caused by the type of primary and secondary infection and its approach in dengue endemic areas. NS1 protein is secreted as a soluble hexamer from DENV-infected cells and circulates in the bloodstream of infected patients.

During the viremic phase of DENV infection, NS1 is concomitantly produced during the virus replication process and is likely to remain in circulation 2 to 3 days after the viremic phase. Antigenemia (NS1) and viremia (RNA) are correlated according to the days of disease (42,43). The validation of diagnostic tests that include NS1 antigen should also be compared with a reference test. In this study, the reference test corresponds to the PCR test through the detection of the DENV genome clinical data and the samples were collected prospectively from patients who were representative of the spectrum of the disease. The results of the rapid diagnosis test and of the reference standard (RT-PCR test) was independently interpreted.

To validate the rapid diagnostic tests that include NS1 antigen seems reasonable considered with as reference a test that measures the same, like ELISA type tests for the detection of NS1 antigens, a comparison that looks as a more equitable. But, if patients with NS1 positive in rapid test have a negative results in the ELISA test the assessment of RDT properties may be biased (differential verification bias) excluding dengue as etiology and, for this way, conducting to a misclassification of cases and invalid data.

When comparing the RT-PCR positivity percentages of 32.9% with that of NS1 of 45.8%, the results could suggest an excess of false positives attributed to the NS1 results of the rapid test. However, molecular biology tests are not 100% sensitive or specific, therefore, and also following CDC recommendations, if a NS1 test is positive, this result confirms a diagnosis of current DENV infection (13). In this study, the overall sensitivity and specificity of NS1 assessed up to 5 and 7 days after symptom onset corresponded to 84.2% and 72.8%; 83.9% and 72.3%, respectively.

In contrast, the results of the viral isolation conducted locally under unblinding conditions, from each sample with a positive NS1 in the rapid test, was obtained a positive result for viral culture. Then, in the context of a dengue outbreak without co-circulation of zika virus, the probability of a result of dengue NS1 false positive is too low.

NS1 protein is secreted as a soluble hexamer from DENV-infected cells and circulates in the bloodstream of infected patients. During the viremic phase of DENV infection, NS1 is concomitantly produced during the virus replication process and is likely to remain in circulation 2 to 3 days after the viremic phase. Important to mention that due to its protein nature it is much more stable than viral RNA molecules, which is why NS1 antigen has become the most common target for detection during an outbreak or epidemic in field work and its detection also allows for rapid results at the point of patient care (44,45).

A differential stability of NS1 antigen and RNA during test processing is suggested as a hypothesis attributing to these differences (44,45). According to our local capabilities for 2019, the rapid tests were run at the same point where the patients were seen, however, the RT-PCR tests were sent to the UVE reference laboratory in France. The samples were aliquoted, as described in the methodology, stored at -80°C locally and then shipped from Bucaramanga to Marseille. This shipment on dry ice could have reached temperatures of -20°C to -40°C during the 7 days of transport and this temperature increase could have affected the integrity of the virus RNA in the samples. Researchers from Brazil found that sensitivity and negative predictive value were directly affected by the preservation conditions of the samples. Tests such as RT-PCR and viral isolation showed a greater dependence on well-preserved samples to make an accurate diagnosis. Their results showed that storage of samples for 15 days at -30°C yielded inaccurate results in RT-PCR and viral isolation tests, while in the same samples the NS1 ELISA tests did not show a significant reduction in positivity (46). In favor of the hypothesis of sample quality deterioration due to transportation, this study shows that the samples that remained at the local level for viral culture processing gave positive results for virus isolation and subsequent genomic sequencing. However, it is important to carry out future studies to replicate this experience and that the processing of samples for all tests be done locally and in the shortest time possible, to reduce sample quality bias.

The non-concordance of RT-PCR and NS1 positivity results attributed to the possible quality of the sample corresponds to a stated limitation of this study. However, showing this weakness of the study also shows a strength and it is to be able to narrate the limitation that molecular biology tests have by requiring referral to specialized laboratories, which for the year 2019 in our region consisted even in making a process of importing samples to another country. The results of the Dengue Duo, were performed on a sample that had less than 2 hours of being collected and where it was run in the same institution and where the patient was being treated, processed by a health professional with a simple training, which did not require additional supplies or reagents and where the result was obtained in less than 8 hours from the time the sample was taken. Thus, this study shows the advantages of POCT for the diagnosis of DENV in a primary health care center in the city of Bucaramanga.

Another important point to discuss about the specificity of this test is to consider cross-reactions of IgM antibodies against other flaviviruses. For the case of Colombia and due to the introduction of ZIKV during 2015-2016, it is important to explore whether this specificity of the IgM of the Dengue Duo test is affected. In this study, molecular biology tests were processed, in which in a single RT-PCR reaction any of the genomic targets of DENV-CHIKV-ZIKV are detected (22,23). The results showed that all samples were positive for DENV and negative for ZIKV and CHIKV.

These results were also compared with the report of the INS/Ministerio de Salud, which showed low circulation of ZIKV and CHIKV in Colombia during the years 2018 to 2020 (36). Based on this and in the clinical context and high prevalence of dengue, in this study the positive IgM result of the rapid tests was considered as a confirmed DENV infection. This suggests that the IgM result could further improve the sensitivity of diagnosis by POCT testing when a patient attends outside of 7 days of symptom onset. It is important to mention that no sensitivity comparison was made in this study comparing rapid test IgM results with IgM detection by ELISA, which is why its use is recommended for future studies.

Diagnostic accuracy depends on the pre-test probability as a function of the prevalence of the disease in the setting. In our study, the prevalence was 32.9%, Positive Predictive Value (PPV) and Negative Predictive Value (NPV) were 60% and 89.9% for NS1. PPV and NPV values reached the highest predictive values at 2 days after the onset of disease symptoms when sensitivity and specificity values are at their highest. From a pragmatic point of view, with a prevalence of 32.9% and on the same day of illness, a clinician who has a rapid test in his office will have a higher positive predictive value for NS1. It is important to clarify that in this study the prevalence of DENV positivity by RT-PCR in this consecutive case series was high; however, it may not reflect the baseline prevalence, given that it has a preliminary clinical assessment. This preliminary clinical evaluation clinically filtered suspicious patients and increased the pretest probability; that is, the clinical examination separated some people from others.

Despite the above scenario of a first medical evaluation and its prior classification, tests that have a high sensitivity also increase their negative predictive values, as in the case of NS1. When the clinician, through the NS1 rapid test, tells a patient that it is negative, even if he suspects it clinically, he has a high probability that it is actually negative. Yow and cols (18) in evaluating six rapid diagnostic tests in Singapore to determine their usefulness in the routine clinical setting for the detection of recent dengue infections found that the PPV and NPV values combining NS1 and IgM were 100% and 90.9%, respectively. However, these results need to be analyzed with caution because they will depend on the prevalence of dengue disease, i.e., talking about an outbreak or epidemic could be very different from periods of low transmission.

Another aspect to be included is the performance of the rapid tests according to DENV serotype. According to the serotyping results in this study, the distribution of serotypes shows that DENV-1 was the predominant serotype (96%) (6), followed by DENV-2 (4%). DENV-3 and DENV-4 serotypes were not detected. These data are consistent with those reported by the INS for the same period in the same region (36). These data correspond to the outbreak and epidemic nature of a single-center study, in which viral circulation can be limited to the detection of circulation of one or two serotypes. Therefore, it could not be determined in this study whether viral serotype could influence the performance of the Bioline Dengue Duo rapid test. Further studies with multicenter design in several geographic regions are required to achieve such representativeness in the same period.

Etiological diagnosis of febrile syndrome was demonstrated by means of rapid tests used at the point of patient care. The sensitization of patients to attend in the early stages of the disease together with their POCT diagnosis, allowed the description of the circulation of DENV serotypes, through the application of molecular biology and viral isolation tests, which even allowed the genomic sequencing of these viruses that circulated during the epidemic of the years 2018 to 2020 in the city of Bucaramanga. Thus contributing to virological surveillance information that could have an impact on the prediction of new outbreaks and epidemics.

It is significant for the interdisciplinary health team that routinely attends to these cases in settings with a high prevalence of dengue and other febrile diseases to have a diagnostic tool that provides results in 30 minutes that can be performed at the point of care of the patient, that is easy to perform and interpret, and that is inexpensive. This strengthens the capacity of first level health centers, improves patient prognosis through timely diagnosis and, in a complementary manner, and contributes to the knowledge of virological surveillance in a region. In dengue endemic areas, the differential diagnosis for acute fever illness involve a broad spectrum of infection including another arbovirus. During the first days of the disease, Dengue and Chikungunya have overlapping symptoms (ie: fever, headache, rash, arthralgia) leading to misdiagnosis. A proportion of dengue cases will develop a life-threatening severe disease (SD). Clinical criteria (warning dengue signs-WDS) are recommended to distinguish between patients who evolve to SD from uncomplicated dengue cases. However, WDS for severe dengue are nonspecific and often develop late in the course of illness. For this reason, in the context of a dengue outbreak, the use in primary health-care services of a dengue rapid test affordable, with high sensitive, specificity and predictive values during the first 2-4 days of the diseases allow provide for patients a close monitoring with the expectation to reduce the risk of severe dengue cases. In conclusion, the validation of a rapid test for NS1 and IgM for the early diagnosis of DENV during an outbreak in Bucaramanga, Colombia was reported and the advantages and limitations of its use in the setting of a primary health care center were shown.

## Data Availability

All relevant data are within the manuscript

## Acknowledgments

We are grateful to Hospital Local del Norte e Instituto de Salud de Bucaramanga. This study was supported by Fundación INFOVIDA and for the European Union’s Horizon 2020 research and innovation program under ZIKAlliance grant agreement no. 734548. RMGR was supported by the French National Research Institute for Sustainable Development (IRD) and Centro de Atención y Diagnóstico de Enfermedades Infecciosas-Fundación INFOVIDA. The funders had no role in study design, data collection and interpretation, or decision to submit the work for publication.

## Author contributions

RMGR and LAV conceptualization. MIE, ATR, MPC and RMGR investigation. RMGR and MPC data curation. LAV, VMH, RMGR methodology. VMH formal analysis. RMGR, LAV, VMH and XDL writing and original draft preparation. RMGR, LAV, XDL, VMH, MIE and MPC writing and review & editing.

